# Efficacy of BianShi Moxibustion for bowel preparation: study protocol for a randomized controlled trial

**DOI:** 10.1101/2024.05.11.24307217

**Authors:** Qiang Zhang, Xiao-Ping Zhang, Li-Jie Wang

## Abstract

**Background:** Adequate bowel preparation is one of the most important prerequisite during the colonoscopy. It is conductive to detect the polypus, adenoma and early colorectal carcinoma and decrease the rate of overuse of medical resource. BianShi Moxibustion(BSM) is one of the most important Traditional Chinese Medicine(TCM) treatment options, which is called TCM proper technology. BSM is applied to prevent and cure digestive system disorder widely, including functional constipation(FC), irritable bowel syndrome(IBS), functional dyspepsia(FD) etc. we speculate that BSM can improve the bowel preparation quality through ameliorating the function of digestive tract, especially in high risk of inadequate bowel preparation patients, including elderly individual, diabetes mellitus, neurodegenerative changes etc. Hence, the objective of this research is to evaluate the efficacy of BSM on the quality of bowel preparation before colonoscopy.

**Method:** This is a randomized triple-blinded, single-center, prospective study. We will recruit 72 inpatients who are scheduled to undergo colonoscopy for screening and diagnostic or therapeutical purposes for the first time will be randomized to assign into treatment group or control group at a ratio of 1:1. The intervention plan in the treatment group consists of 3L polyethylene glycol(PEG) solution plus BSM(Treatment period is 3 days, Qd). In the control group, the plan is 3L PEG plus sham BSM. Boston Bowel Preparation Scale(BBPS) will be used to evaluate the efficacy of the bowel preparation and regarded as the primary outcome measure. The secondary outcomes including caecal intubation rate, the willingness to repeat colonoscopy, the tolerance of bowel preparation regimens, the rate of adverse events.

**Discussion:** The aim of this clinical trial is to confirm the influence of BSM on the quality of bowel preparation. The hypothesis of this study is that the BSM has a positive role on the quality of bowel preparation. The routine bowel preparation regime combines with TCM proper technology will enhance bowel preparation quality and ameliorate the subjective feeling of participants. In addition, it will provide the reliable evidence of bowel preparation prior to colonoscopy from the point of view of integrative medicine.

**Trial registration:** This protocol and trial was registered in the Chinese Clinical Trial Registry(ChiCTR2300077168).

## Introduction

Colonoscopy is regarded as one of the most widely used tools to screen the colorectal disorders with diagnostic and therapeutical potential. Adequate colonoscopy is pivotal to detect polys, adenoma, precancerous lesion in bowel. With the advent of the aging society, the rate of adenomatous polyps with precancerous potential is increasing. Hence, colonoscopy has already become one of the major routine detection tools in the large-bowel and terminal ileal disease. Many factors have an impact on the quality of colonoscopy. Whereas, adequate bowel preparation is a prerequisite for colonoscopy. Inadequate bowel preparation not only decrease the detection rate of polyps, increase the risk of suffering precancerous lesion of bowel and the risk of rechecking, but also prolong the incubation time. Therefore, eligible bowel preparation is of pivotal importance for colonoscopy. According to the relevant research, 20%-30% of patients without obtaining adequate bowel preparation before colonoscopy^[1].^ Some studies indicated that long-term conspitation, diabetes mellitus, Parkinson disease, poor compliance, elder et al are risk factors for inadequate bowel preparation^[2-5]^. In some circumstance, those risk factors reflect the poor gastrointestinal function of patients. The relative poor gastrointestinal function not only are related with inadequate bowel preparation, but also related with the adverse events, such as nausea, emesis, bloat, abdominal pain et al. Therefore, the definition of excellent bowel preparation project is safe, effective, meanwhile being well-tolerated.

BSM is one of the therapeutical technologies in TCM, which combines the Tuina and Moxibustion. It is widely used to treat multidisciplinary illness, especially in the spectrum of alimentary system, such as functional constipation(FC), irritable bowel syndrome(IBS), functional dyspepsia(FD) et al. BSM is a special medical device made of microcrystalline limestone, and the interior is filled with burning artemisia, a kind of traditional herb in China. The details of manipulation is to press on the abdomen with this special medical device in the clockwise manner, stimulate the special acupoint on the abdomen, and then rub and press the abdomen in the way of special Tuina to relax the local tissue. We speculate that the curative power on abdominal diseases of BSM is conductive to regulate the bowel movement, improve the local circulation, finally, being helpful to enhance the quality bowel preparation and prevent the occurrence of uncomfortable symptom during the period of bowel preparation. Hence, we design this randomized multi-blinded, controlled, clinical trial to evaluate the efficacy of BSM on the quality of bowel cleaning.

## Objective

This randomized controlled trial aims to (a)evaluate the potential of BSM on the quality of bowel preparation before colonoscopy. (b)evaluate the effect of BSM on reducing the adverse symptoms during the period of bowel preparation and ameliorating the subjective feeling of the patients.

## Method/design

This research is a prospective, single-center, randomized, controlled, multi-blind(colonoscopist, statistician, patient) clinical study, and it will be conducted from November 2023 to November 2024. The eligible patients will be divided into treatment group and controlled group at a ratio of 1:1 randomly. The randomization sequence will be generated via the software SPSS.19.0 and inform the attending physician through the independent staff. The patients in the treatment group will receive 3-day BSM treatment(once daily) period, meanwhile, the patients in the controlled group will get 3-day sham-BSM(once daily) treatment period. The colonoscopy of all the participants will conducted in the morning. The flowchart is shown in Fig.1. This trial will be conducted in line with the standard Protocol Items Recommendations for Interventional Trials(SPIRIT) guidelines^[6]^ and written informed consent will be obtained from all the participants. All the procedures in this trial will adhere to the principles of the Declaration of Helsinki(Version 2000). This clinical trial has already registered in the Chinese Clinical Trial Registry(registry number: ChiCTR2300077168) and approved by the hospital ethics committee of ShenZhen hospital of BeiJing university of Chinese medicine(Long Gang).

**Fig. 1.**
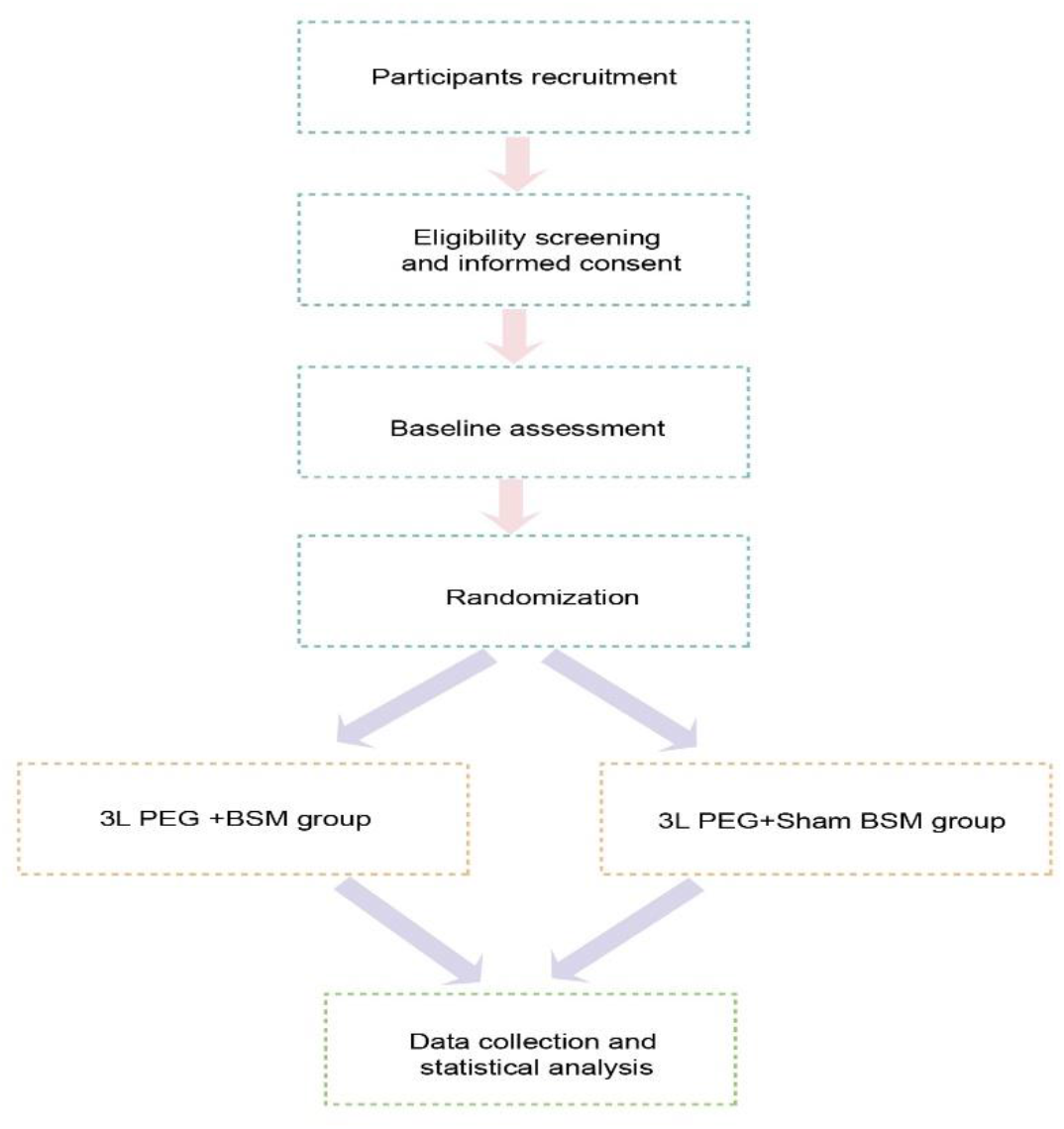
Flowchart of study design.

### Participant

This clinical trial will recruit a total of 72 inpatients in the on-site way. All the participants will come from ShenZhen hospital, BeiJing University of Traditional Chinese Medicine(Long Gang). All the participants have the right to withdraw at any time and for any reason. In addition, the treatment regimen of all the participants will not be affected because of withdrawal.

### Inclusion Criteria

1. Participant aged between 18-75;
2. Be able to adhere to medical order to finish the procedures of bowel preparation;
3. Participants who obtain a complete colonoscopy;
4. Participants who agree to join and write informed consent;

### Exclusion Criteria

1. Participants who are allergic to any component of PEG;
2. Participants with poor health, including heart failure, cirrhosis, chronic kidney disease or with clotting mechanism disorder;
3. The absolute contraindication of colonoscopy including obstruction, perforation, severe inflammatory bowel disease;
4. Participants with a history of previous abdominal surgery;
5. Participants suffer from mental disorder, meanwhile, cannot adhere to medical orders;

### Elimination Criterion

1. Participants who cannot fill fully out the questionnaire;
2. Various reasons which lead to a incomplete colonoscopy;
3. Participants who cannot complete the PEG-based bowel preparation regimen or not complete the treatment period of BSM;
4. Severe advent events is present during the period of bowel preparation;

### Sample Size Calculation

The sample size was calculated based on the results of pre-clinical trial. The estimation formula as follows: N_1_ = N_2_ = 2 × (Z_α + Z_β)^2 × σ^2/(μ_1 - μ_2)^2 two-sided test w s used. Based on the statistic l results from our pilot study μ_1_ in the treatment group is 8.07 and μ_2_ in the control group is 6.93 respectively. The standard error *σ*=1.417. We set the power of test 1-β=90% namely β=0.1 and type I error rate α=0.05. Thus the calculated sample size of each group is approximately 32 cases. Considering the dropout rate of 10% is allowed hence the tot l number of 72 patients re required to be included in this tri l.

### Randomization, Blinding, Implementation

Participants which are eligible to the inclusion criterion will be randomly assigned to the treatment group or control group at a ratio of 1:1. Random sequences will be generated through SPSS version 22.0 software. The number of random sequences will be delivered by independent member in the way of opaque envelope to the attending physicians. Endoscopic physicians, data analysists, participants in this trial will be blinded in the manner of masking private information of participants in order to eliminate the bias as much as possible. During the procedures of this trial, all the relevant researches have the right to carry out unblinding in confront of severe advent events, including burns, exacerbation of pre-exist conditions or participants request for withdraw..

The bowel preparation regimen in the treatment group of this trial including 3 pockets of PEG(specification:68.56g) plus BSM treatment (period:3 days). The details as follow: the patients will be instructed to administer 1 bag PEG solution(volume:1L, temperature:38 °C) in one hour at 8:00 pm before colonoscopy. The second PEG solution(volume:1L, temperature:38°C) will be taken at 4 am in the day of colonoscopy. The third PEG solution(volume:1L, temperature:38°C) will be taken at 6 am in the day of colonoscopy. Every 250ml PEG solution will be completed in 15 minutes in case of uncomfortable symptoms such as nausea, vomiting, abdominal pain etc. All the colonoscopy will proceed in the morning.

The implementation details of BSM as follows:

1. Daubing abdomen with essential oil;
2. Kneading abdomen in the manner of clockwise with both hands 36 times;
3. Kneading the Dai meridian from the origin to the terminal point 36 times;
4. Pressing on the acupoint of Shenque(RN8) with BSM device(columnar and burning wormwood inside of this device) lasting one minute;
5. Kneading the Ren meridian from above down in the abdominal region with BSM device 10 times;
6. Kneading the stomach meridian which belongs to abdomen region from above down with BSM device 10 times;
7. Stimulate the acupoints including Zhongwan(RN12), bilateral Tianshu(ST25), Qihai(RN6), Guanyuan(RN4) with BSM in the manner of clockwise 10 times;
8. Clean the regional skin and keep warm;
9. Pay attention to the strength during the conduction of BSM.

The same manipulation of BSM will be conducted on the abdomen in the control group(sham group). The key difference point is (1)First of all, there is no burning artemisia inside of the BSM device; (2)The second difference point is the manipulation strength will not result in the shape change of the abdomen.

The diagrammatic sketch of BianShi Moxibustion device is presented in Fig.2.

**Figure 2.**
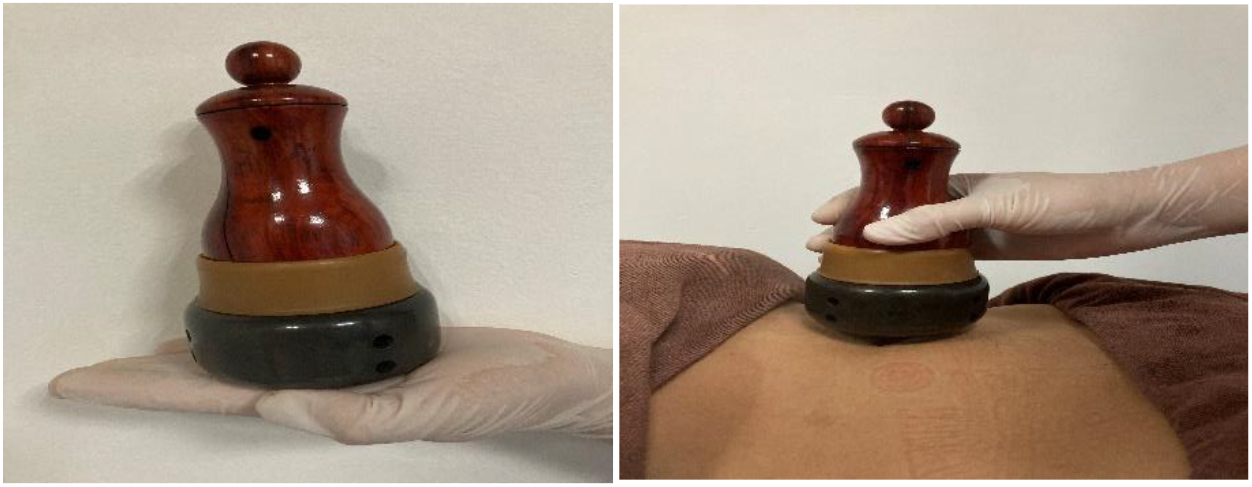
BianShi Moxibustion device.

### Primary outcome

We will regard the quality of bowel preparation as the primary outcome. We will utilize the Boston Bowel Preparation Scale(BBPS) to evaluate the quality of bowel preparation. For more details of specific evaluation method, see supplement 1. The total score ranges from 0 to 9 point and the score of every segment of bowel including left, transverse, right colon ranges from 0 to 3. The score of every segment bowel ≥2 point, meanwhile, the total score ≥6 point is considered to be eligible. The higher score manifests the better quality of bowel preparation.

### Secondary outcomes

Secondary outcomes including the interval between the start of bowel preparation and the first bowel movement, total number of bowel movement, tolerability of bowel preparation, incidence rate of adverse events including abdominal pain, bloating, nausea, vomiting.

### Data collection and management

All the researchers will be required to sign confidentiality agreement to protect the participants’ private information and privacy as much as possible. Case Report Form(CRF) will be recorded in the manner of Excel chart. The items of CRF including gender, age, body mass index(BMI), history of drinking and smoking, level of education, concomitant diseases including chronic constipation, Parkinson’disease, history of stroke or spinal cord injury, use of tricyclic antidepressants or narcotics, inflammatory bowel disease(IBD), diabetes, chronic liver disease.

### Surveillance of data

In order to ensure the integrity of the clinical data, we will conduct data collection once week by the specific statistician to achieve the goal of integrity of data and record surveillance.

### Statistical analysis

After the clinical data being collected, categorical variables will be presented as frequencies and percentages. Continuous variables will be expressed as the mean± standard(SD). The Fish’s exact test and two-sided test will be used when analysing the categorical data and continuous data respectively. SPSS 19.0 software(IBM Co. Armonk, NY, USA) will be utilized for statistical analysis. The significant criteria will be set at *p*<0.05.

## Discussion

Colonoscopy is one of the most widely used technologies to diagnose and treat bowel-related diseases. The incidence of intestinal lesions such as polyps, adenomas, early colorectal carcinoma etc has a increasing tendency with age, especially in the aged society of China. Complete colonoscopy and clearly view is significant to evaluate the lesion of bowel objectively. Obviously, adequate bowel preparation is one of the vital procedures for the colonoscopy.

Functional constipation, diabetes, neurodegenerative lesions, old age etc are risk factors for inadequate bowel preparation and those risk factors are related to the decreased gastrointestinal function. Hence, designing safe and effective bowel preparation protocol before colonoscopy is necessary to improve the detection rate of intestinal diseases. BSM, as one of the proper techniques of Chinese medicine, is widely used in managing diseases of several clinical disciplines, such as gastroenterology. BSM stimulates the special regions, meridians or acupoints of the body in the way of combining moxibustion and Tuina subtlely to improve the symptoms and cure the diseases. Some researches indicates that techniques of TCM, such as acupuncture, moxibustion and Tuina has a positive efficacy on ameliorating the symptoms of functional gastrointestinal disorder including IBS, FC, FD^[7-9]^.

BSM has the technical characteristics and advantages of moxibustion and massage. While, to the best of our knowledge, there is no relevant clinical research to evaluate the efficacy of BSM on the quality of bowel preparation. Hence, we design a randomly, controlled, multi-blinded clinical trial to validate the effectiveness of BSM on the cleanness of bowel, meanwhile, we will demonstrate the safety of BSM, the tolerance of bowel preparation program. The “Chinese Guideline for Bowel Preparation for Colonoscopy”(2019) only recommends senna, a widely used herb to manage constipation, as an integrative medicine regimen for bowel preparation before colonoscopy, which restricts largely the application and development of complementary and alternative medicine. Therefore, it is necessary to explore further the potentially clinical value of proper techniques of TCM.

Although, this study adhere strictly to the principles of RCT trial, several limitations still exist. First, this is a single-center clinical study, which will degrade the level of evidence to some extent. Second, the effectiveness of blinding is not evaluated in the sham BSM group, which probably produce the confounders. Based on the mentioned limitation above, the further study will overcome those defects.

In conclusion, this is the first RCT study with high-quality to certify the effectiveness, safety, and advantage of BSM. The results will contribute to the development of comprehensive bowel preparation protocol in the perspective of evidence-based medicine.

## Data Availability

All data produced in the present study are available upon reasonable request to the authors

## Contributors

QZ is responsible for designing the trial, drafting the protocol. XPZ is responsible for the methodology of the trial. LJW is accountable for all aspects of the work. All the authors read and approved the final manuscript of this prototol.

